# Investigation into Quality of Life and Psychological Status of Different Populations during COVID-19: A study concerning Surrounding Areas of Wuhan

**DOI:** 10.1101/2020.08.15.20158725

**Authors:** Zheng Liu, Jingsong Mu, Wenxiang Fan

**Affiliations:** Department of Rehabilitation Medicine, The First Affiliated Hospital of USTC; Division of Life Sciences and Medicine, University of Science and Technology of China, Anhui Provincial Hospital, Hefei, 230001, China

**Author notes:** Zheng Liu, Corresponding author: Jingsong Mu, Wenxiang Fan.

**Keywords:** Coronavirus Disease 2019 (COVID-19), Quality of Life (QOL), Infectious Disease Outbreaks

## Abstract

**Objective:** To investigate different populations’ quality of life and psychological status in surrounding areas of Wuhan during COVID-19 pandemic.

**Methods:** The data of 248 residents living in Anhui from February 4 to 6 of 2020 were collected through network surveys including age, gender, occupation, the World Health Organization Quality of Life measurement Scale short form (World Health Organization Quality of Life instrument brief, WHOQOL BREF), Zung Self-rating Anxiety Scale (Self-rating Anxiety Scale, SAS and Zung Self-Rating Depression Scale (SDS). Those surveyed, divided into two groups: medical staff (129 cases) and nonmedical staff (119 cases), were made statistic analysis according to the factors mentioned above.

**Results:** The WHOQOL-BREF of medical staff in this region was lower than that of nonmedical staff in the fields of physiology, psychology, social relations, and environment, among whom female medical staff scored significantly lower than that of male medical staff in four fields. There was no significant statistical difference in SAS and SDS scores between the two groups, and gender had no significant influence on SAS and SDS scores of medical staff.

**Conclusion:** During the COVID-19 pandemic, medical staff enjoyed a lower quality of life in surrounding areas of Wuhan than that of nonmedical staff, and female medical staff even lower, which should arouse social concerns.

## Introduction

COVID-19 tends to seriously threaten the health and life of people. Some studies have been made on working status, quality of life and psychological status of medical staff in the harder-hit areas like Wuhan^[1]^, while in the already infected surrounding areas, medical staff face much higher pressure than before due to dense population and mobilization of local medical resources to Wuhan. Related studies on that population appear to be helpful for pandemic response in other areas. Therefore, medical and nonmedical personnel in Anhui province, adjacent to Hubei Province, were selected for this study.

### Data Sources

Collect data of 248 residents living in Anhui from February 4 to 6 of 2020 through a Network questionnaire survey, among whom 146 are male, 102 are female, ranging from 18 to 75 years old. They are divided into the medical staff group (129 cases) and the nonmedical staff group (119 cases) whose general data are shown in table 1. In the two aspects of gender and age, there was no statistically significant difference (P > 0.05).

**Table 1.** comparison of general data between the two groups

| Group | n | gender |  | Average age<br>(year) |
| --- | --- | --- | --- | --- |
|  |  | Male | Female |  |
| medical staff | 129 | 75 | 54 | 33.79±9.906 |
| Nonmedical<br>staff | 119 | 71 | 48 | 36.74±8.836 |

## Methods

Before the questionnaire survey, 268 residents were informed and agreed to cooperate, of which 248 residents completed the questionnaire as required. They were conscious and had no serious cognitive impairment when filling in the questionnaire. The questionnaire included gender, age, occupation, Chinese version of World Health Organization Quality of Life instrument brief (WHOQOL-BREF), self-rating Anxiety Scale (SAS), self-rating Depression Scale (SDS), etc.^[2-4]^ The standardized full score of each field is 100.

1. The Chinese version of WHOQOL-BREF quality of life measurement scale can generate scores in four fields, physiological, psychological, social relations, and environmental fields. The field score was recorded in a positive way, that is, the higher the score, the better the quality of life.
2. The anxiety self-rating scale (SAS) is used to assess the subjective feelings of anxious people. It consists of 20 items including 15 items of positive score and 5 items of negative score. The standard threshold was 50 points, with 50-59 being considered mild anxiety, 60-69 moderate anxiety, and 69 above severe anxiety.
3. The self-rating depression scale (SDS) is used to assess the subjective feelings of depressed people, including 20 items. The scoring method is the same as the anxiety score. The cut-off value of SDS standard score was 53, among which 53-62 were classified as mild depression, 63-72 as moderate depression, and 72 or more as severe depression.

## Statistical Methods

SPSS 17.0 statistical software was used for data analysis. Independent sample t-test was used for comparison of measurement data conforming to normal distribution, while rank sum test was used for comparison of measurement data not conforming to normal distribution.Chi-square test was used to compare counting data, and P < 0.05 was considered statistically significant.

## Results

### 1. Comparison of scores of each scale between the two groups

The scores of Zung self-rating scale for depression and Zung self-rating scale for anxiety in the two groups were in line with the normal distribution. The t-test results indicated that there was no statistically significant difference in the scores of Zung self-rating scale for depression and anxiety in the two groups. Rank sum test was used for the scores of physiological field, psychological field, social relationship field, and environmental field. The results indicated that the scores of medical staff were lower than those of nonmedical staff in the four fields, as shown in table 2.

**Table 2.** comparison of survey results among different groups (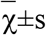)

| Group | Physical field | Psychological field | Field of social relations | Environment field | SAS standard score | SDS standard score |
| --- | --- | --- | --- | --- | --- | --- |
| medical staff<br>(n=129) | 78.693±8.191 | 70.930±7.488 | 75.401±8.490 | 70.388±6.199 | 41.550±9.226 | 43.866±10.916 |
| Nonmedical staff(n=119) | 84.106±5.600 | 80.000±4.593 | 79.384±3.782 | 73.929±3.633 | 40.893±8.387 | 42.658±11.303 |
| p-value | <0.001 | <0.001 | <0.001 | <0.001 | 0.559 | 0.393 |

### 2. Comparison of scores of various scales among medical staff of different genders

The scores of Zung self-rating scale for depression and Zung self-rating scale for anxiety in the two groups were in line with the normal distribution. The t-test results indicated that there was no statistically significant difference in the scores of Zung self-rating scale for depression and anxiety in the two groups.Rank sum test was used for the scores of physiological field, psychological field, social relationship field, and environmental field. The results indicated that the scores of female medical staff were lower than those of male medical staff in the four fields, as shown in table 3.

**Table 3.**
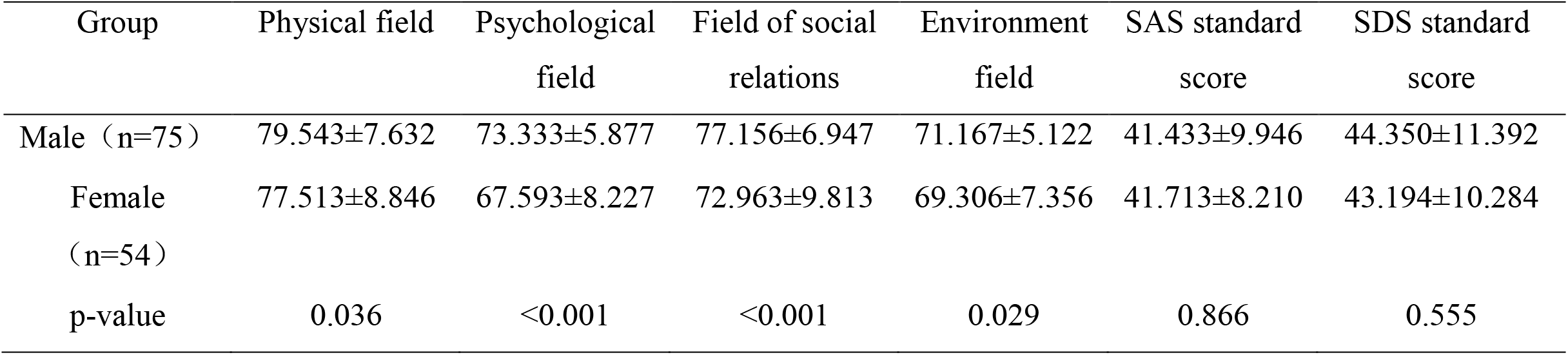
comparison of findings between medical staff of different genders (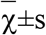)

### 3. Comparison of scores of various scales among nonmedical staff of different genders

The scores of Zung self-rating scale for depression and Zung self-rating scale for anxiety in the two groups were in line with the normal distribution. The t-test results indicated that there was no statistically significant difference in the scores of Zung self-rating scale for depression and anxiety in the two groups. Physiological, psychological, social relations, and environmental score was reached by the rank sum test, of which the results indicate that nonmedical staff of different genders in the quality of life scores had no significant statistical significance, but female nonmedical staff Zung depression scale score seems higher than men with the p-value 0.097, as shown in table 4.

**Table 4.**
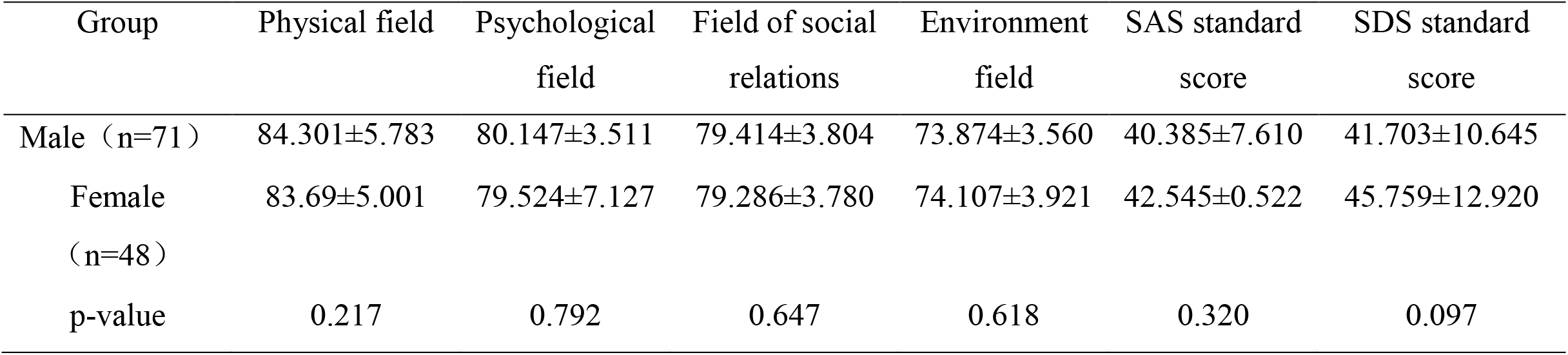
comparison of findings for nonmedical staff of different genders (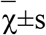)

## Discussion

In terms of the impact of public emergencies on the quality of life of the population, previous studies have suggested that different groups are affected differently, and most of them have shown that public emergencies have adverse effects on medical staff, including the decline in the quality of life. ^[1,5,6]^ This study found that the score of WHOQOL-BREF of medical staff was lower than that of nonmedical staff in physiological, psychological, social relationship, and environmental fields, which was similar to most previous research results. However, Cornelia studies show that these incidents do not have much impact on people’s quality of life and the impact is often transient.^[7]^ Perhaps because studies of Cornelia et al. focused on noncommunicable events in which the studying objects were not subjected to extensive isolation for long periods of time. Cong et al. showed that the change of living order and habits during the SARS outbreak in isolation may lead to the decline of life quality and psychological state. ^[8]^ During this outbreak, to control the spread of the disease, the government required most people to cancel their work and stay in quarantine at home, while medical staff were responsible for the prevention and control of the epidemic. All these factors may further reduce the quality of life of medical staff.

Previous studies have shown that public emergencies have different effects on medical staff of different genders. The majority of previous studies believe that public emergencies have a greater impact on women’s quality of life than men’s. ^[9-11]^ Our results suggest that women’s quality of life scores were lower than men’s in all four areas among medical staff, while there was no statistically significant difference in quality of life between the two genders among nonmedical staff. Previous studies by Mariza and Sergio et al. have shown that overwork, lack of external support, and poor sleep quality are important reasons for the decline of people’s quality of life. ^[12]^ COVID-19 happened to occur in China’s traditional Spring Festival. Such pandemic forced medical staff to cancel the vacation and dedicate to the prevention and treatment work. Fatigue may lead to a decline in their quality of life. And the situation of women may be more visible than that of men. At the same time, the outbreak has extended the leave of nonmedical staff, whose quality of life scores may be higher due to the presence of family members. In addition, there were no adverse factors such as overwork, lack of family support, and poor sleep quality among nonmedical staff.So men’s physical advantages may not show up. As a result, the difference in quality of life between men and women is not significant among nonmedical staff.

In terms of psychological state, previous studies based on the outbreak of SARS and MERS have shown that public emergencies can lead to an increased risk of anxiety and depression. ^[13]^ For example, Angelina et al. found that a considerable number of medical staff were affected by emotional and mental trauma during SARS, and pointed out that clear and effective preventive measures and support from superiors and colleagues were important protective factors. Imran et al. showed that during the MERS outbreak, the mood of medical staff was generally affected and pointed out that the clinical improvement of infected colleagues and the blocking of cross-transmission after taking strict protective measures all alleviated their bad mood. However, there were still some different voices. For example, the study of Wang showed that the medical staff of SARS in wuhan region did not have obvious psychological emotions of anxiety and depression on the whole, but female medical staff may be more prone to anxiety and depression than men in terms of psychological changes. ^[14]^ The results are similar to ours. We think the possible reason that lead to the results are: 1. Angelina and Imran’s research object is the outbreak of the earlier people, respectively, 2 months after the SARS outbreak in China, 1 month after the MERS outbreak in Arab. Early in an outbreak, when people know less about new outbreaks, they are prone to changes in their psychological states. Wang’s study was conducted four to five months after the SARS outbreak, and at that time people had a certain understanding of SARS; 2. Although our study was carried out in the early stages of the outbreak, this time the situation was different. After the outbreak of the pandemic, the government and medical institutions promptly organized medical staff to study and strengthened the publicity of knowledge, as previous studies have shown that clear and effective learning measures can alleviate bad emotions. ^[11]^The rapid popularization of relevant knowledge reduced the bad mood; 3. The fatality rate of this COVID-19 is far less than that of SARS and MERS. According to recent research results, it is believed to be around 1.4. ^[15]^ In addition, the subjects of this study were mainly concentrated in areas outside Hubei, which made the psychological impact caused by the pandemic smaller. However, it is worth noting that the SDS standard score of women in nonmedical staff has a higher trend than that of men (P=0.097), and it is still necessary to pay attention to women in this group.

In summary, during the outbreak of COVID 19, different populations were affected differently. Due to the particularity of work, the quality of life of medical staff in some fields is low, among which the negative impact on female medical staff is more prominent. Timely support and concern may be more conducive to helping medical staff on these areas better face the challenges of the pandemic.

## Data Availability

All data generated or analyzed during this study are presented in the manuscript. Please contact the first author for access to the data presented in this study.

## Acknowledgment

We thank the hospital staff for their efforts in this investigation, and network staff for their dedication to data entry and verification.

